# Knowledge and awareness of Dengue and Chikungunya amidst recurrent outbreaks amongst urban slum community members of Pune, India

**DOI:** 10.1101/2020.05.14.20101394

**Authors:** Swapnil R. Godbharle, Bibek R. Giri, Abhay M. Kudale

**Affiliations:** Health Sciences, Interdisciplinary School of Health Sciences, Savitribai Phule Pune University, Pune; Master of Public Health, Interdisciplinary School of Health Sciences, Savitribai Phule Pune University, Pune; Interdisciplinary School of Health Sciences, Savitribai Phule Pune University, Pune

**Keywords:** Dengue, Chikungunya, Knowledge, Public Awareness, WHO GVCR

## Abstract

**Context:** World health organization’s (WHO) new strategy on Global Vector Control Response (GVCR) considered engaging and mobilizing communities and emphasized on local adaptation of communication strategies for achieving sustainable impact in vector control. Public awareness studies amidst and post epidemic/ outbreak situations can fast-track useful information to local programmes to set risk communication agenda.

**Aims:** To assess knowledge and awareness regarding dengue and chikungunya among general community members during and post-epidemic/outbreak situations.

**Settings and Design:** Cross-sectional study in slums of Pune city wherein several recurrent outbreaks of Dengue and Chikungunya were reported.

**Methods and Material:** In total 309 general community members residing in urban slums of Pune in Maharashtra were randomly chosen using population proportionate sampling. Semi-structured interview schedule was administered.

**Statistical analysis used:** Knowledge and awareness scores for dengue and chikungunya were calculated and analytical statistical tests were performed.

**Results:** It was found that most of the respondents were in the 18–39 years age group, ever married, literate, and had regular income. The study showed relatively good knowledge among respondents about symptoms, causes, mode of transmission and prevention methods for dengue and chikungunya. Respondents scored well with regard to awareness indicators concerning availability of supportive and symptomatic treatments for dengue and chikungunya. Average knowledge and awareness score for dengue and chikungunya together were 5.4 ± 1.96 (SD) and 1.68 ± 0.63 (SD) respectively. Better knowledge and awareness levels were reported among literate respondents.

**Conclusions:** Findings indicate need for sustaining these better knowledge and awareness levels into efficacious and community-owned prevention practices to fast-track appropriate public health actions during and post-epidemic situations leading to sustained community mobilization and engagement to promote successful implementation of WHO recommended GVCR strategy.

## Background

Worldwide social, demographic and environmental factors strongly influence transmission patterns of vector-borne pathogens, with major outbreaks of vector borne diseases such as Dengue, Chikungunya and Zika virus disease since 2014^1,2^. In view of these ongoing VBD outbreaks in many countries in 2017, the World Health Organization has developed a new strategy to strengthen vector control globally, widely regarded as the Global vector control response 2017–2030 (GVCR). The 70th World Health Assembly adopted resolution of GVCR and encouraged Member States to develop or adapt national vector control strategies and operational plans in consonance with this global strategy. Engaging and mobilizing communities has been considered as one of the important pillars of action in GVCR strategy. And in spelt out research needs GVCR document emphasized on adaptation of information, education and communication strategies to ensure acceptability, participation and appropriate use of vector control tools^1,2^.

Vector control has been considered worldwide as one of the best ways to control any VBDs. The success of vector control methods depends on public engagement and pro-active social mobilization. It is important to assess public awareness regarding the VBDs, along with its mode of transmission and prevention measures^3^. Public awareness studies can serve as a useful tool to strengthen vector control response and necessary public health actions^4^. This information can help the programmes to set communication agenda linked to increased public engagement and mobilizing demand for necessary services. It can also help in developing tailored strategies which can be appropriate for the socio-cultural and politico-economic contexts of different communities^2^. Pune city in Maharashtra India had been hit by several Dengue and Chikungunya outbreaks in recent years^3^. The poor environmental and sanitation conditions in the slums not only contribute to the spread of the disease but also make it difficult for local municipal corporation health services to control the vector population effectively in these areas. Hence, a cross-sectional study was carried out to assess the knowledge and awareness regarding Dengue and Chikungunya among general community members residing in slums of Pune city.

## Subjects and Methods

### Study setting

Pune is the second largest city in the Indian state of Maharashtra, after Mumbai. It is the ninth most populous city in the country with an estimated population of 3.13 million^5^. Along with its extended city limits Pimpri Chinchwad and the three cantonment towns of Pune, Khadki and Dehu Road, Pune forms the urban core of the Pune Metropolitan Region (PMR)^6^. According to the 2011 census, the urban area has a combined population of 5.05 million while the population of the metropolitan region is estimated at 7.27 million^7^.

A community-based cross-sectional study was carried out during 2017–18 in urban slums which were low-income settings in Pune Municipal Corporation (PMC), Maharashtra, India. The sampling method was driven by the need to include a representative number of participants for the study purposes. The results of previous knowledge and awareness studies on Dengue and Chikungunya in India were reviewed taking into consideration key areas of enquiry and indicators to be examined in the present study^8–10^. The review provided the expected minimum value of knowledge and awareness proportion as 0.25. The formula for calculating sample size for proportions of large populations was used. One-third of the total 15 administrative wards in Pune city (5 administrative wards) were randomly selected for the survey. Population proportionate sampling was used to calculate a sample size of 309 respondents from 5 administrative wards. From each of the selected wards one low-income setting officially regarded as slum having highest number of slum structures was selected. Altogether a total of five slums (low-income settings) were selected for the study Among the general community member participants from each slum a 1:1 sex ratio was maintained. Based on the number of slum structures of five selected slums, the population proportionate sampling for each respective slum was calculated. The households from every selected slum were randomly selected and one adult member from each household was interviewed if fitting into the pre-decided inclusion criteria. Inclusion criteria were community members who were residents of PMC (Pune Municipal Corporation) area and aged between 18–60 years.

A semi-structured pre-tested interview schedule was administered through tablet version of “EpiCollect 5” for data collection by trained investigators^11^. All the questions asked in the interview schedule were multiple choice questions. Pilot testing was done before conducting the main survey study. For validation of tool, the interview schedule was checked for the flow and consistency of questions, and then it was revised which mainly comprised of formatting changes. The survey helped in documenting public knowledge and awareness about Dengue and Chikungunya symptoms, causes, modes of transmission, and prevention methods, as well as awareness about availability of the treatment. The complete data collection process was closely monitored by research supervisor to ensure overall quality control. Written informed consent was obtained from individual general community member prior to the interview. The entire study protocol was approved by institutional ethics committee of Savitribai Phule Pune University (Ref. SPPU/ISHS/117/2018).

Survey data were analysed into SPSS (Statistical Package for the Social Sciences, Chicago, IL, USA), version 19. Descriptive statistics were initially performed to summarize data, and the data were categorized into knowledge and awareness indicators to calculate the scores for each category. Knowledge indicators were those providing biomedical information or facts about Dengue and Chikungunya, while awareness indicators were related to the availability of treatment services and hearing information about Dengue and Chikungunya. The Chi-square test was performed to determine the factors associated with socio-demographic indicators such as gender, age, education and marital status. The t-test was used to study association between knowledge and awareness indicators across education. For all analysis purposes, p value less than 0.05 (two-tailed) was considered for statistical significance.

### Development of Dengue and Chikungunya Composite knowledge and awareness scores

Descriptive statistics were initially performed to summarize data, and the data were categorized into knowledge and awareness indicators to calculate the scores for each category. After testing correlation between items for each knowledge topic, 9 knowledge items were retained as valid and included in the calculation of knowledge score as follows: for each individual item, correct answer recoded into ‘1’ and ‘incorrect or don’t know’ answers into ‘0’. A single summary score for each knowledge topic was obtained by summing the number of correct responses and dividing the total by the number of items in the topic. An aggregate measure was then computed, summing the values of each topic yielding a knowledge score for each participant. The theoretical range of knowledge was 0–9, with a higher score indicating a higher level of knowledge. The average knowledge-scores for the group were obtained by summing each participant’s score and dividing it by the total number of survey participants.

### Calculation of maximal mean knowledge and awareness score

Mean knowledge and awareness score by topic as percentages of maximal scores were calculated and considered as maximal mean knowledge and awareness scores. For mean knowledge score maximal theoretical possible score was 9 and for mean awareness score maximal theoretical possible score was 2.

## Results

Out of the total respondents 50% (154) were men and 50% (155) were women. The mean age of respondents was 32 ± 10.77 (SD). More than half (n = 231, 75%) of the total 309 respondents were in the younger age group (18–39). Of total, almost three-fourth of the respondents (n=229, 74%) were under the category of ever married and rest were in never married category. Among the respondents, 91% (n=280) respondents were literate, that means they have at least attended formal education. Table 1 shows the socio-demographic characteristics of the respondents.

**Table 1:**
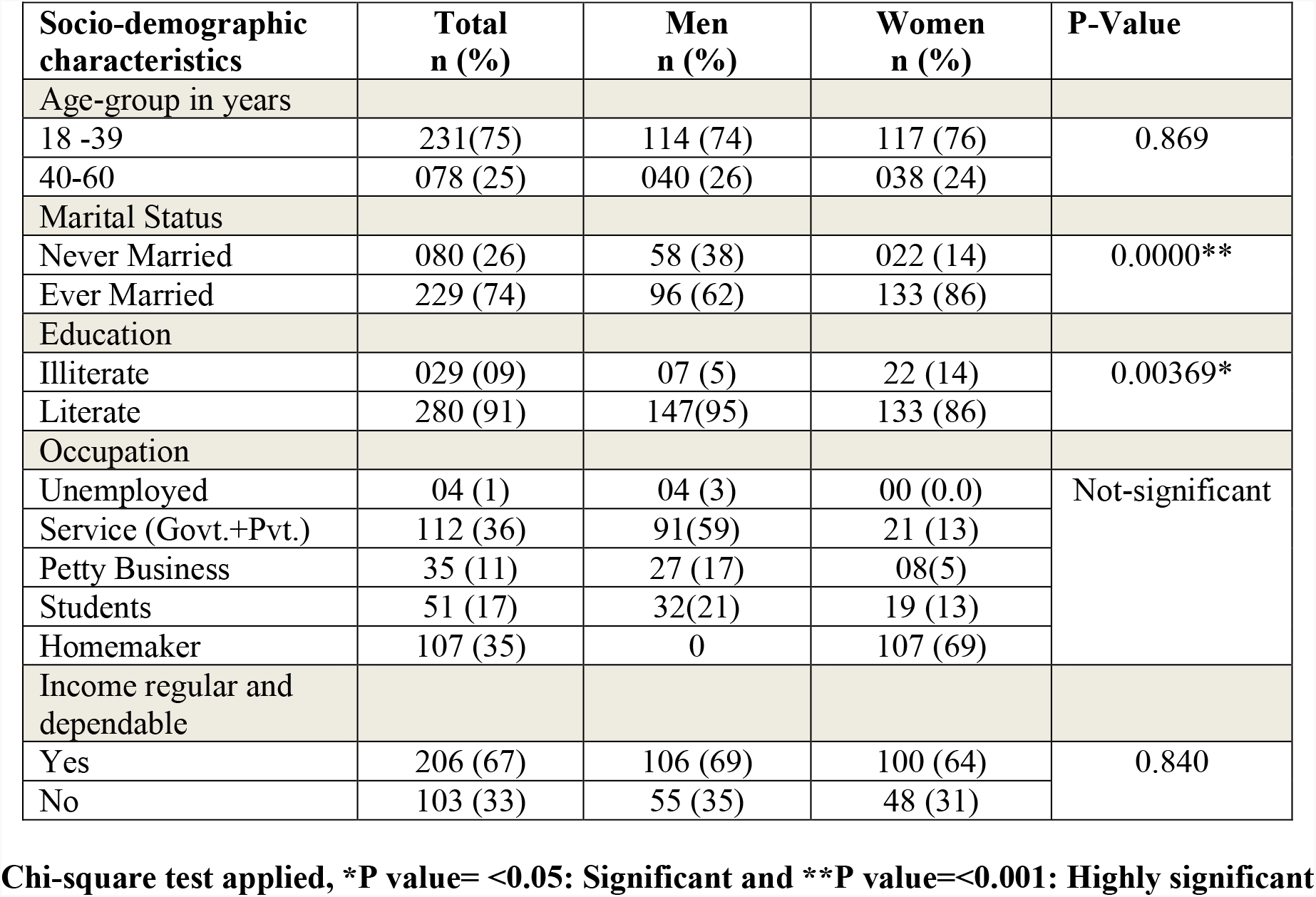
Socio-demographic profile of the respondents (n = 309)

### Chi-square test applied, *P value = <0.05: Significant and **P value = <0.001: Highly significant

Table 2 shows the respondents knowledge about Dengue and Chikungunya. For Dengue high fever(76%) was the most commonly reported symptom, followed by nausea and vomiting (34%), while for

Chikungunya joint swelling (65%) was the most commonly reported symptom, followed by high fever (32%). More than one-third of the respondents (44%) reported virus as a cause for Dengue and two-third of the respondents (66%) were able to report virus as a cause for Chikungunya. More than two-thirds (70%) reported mosquito bite as a mode of transmission of Dengue whereas more than three-fourth of the respondents (78%) were able to report mosquito bite as a mode of transmission for Chikungunya. Prevent standing/ stagnant water near house was reported as a common way of preventing Dengue by 81% (n = 250) of the respondents, followed by Indoor residual spray (71%). Almost 90% of the respondents were able to report that prevention methods of Chikungunya are similar to those of Dengue. There was no statistically significant gender-differences across knowledge indicators of Dengue and Chikungunya. Respondents knowledge about Dengue and Chikungunya was found to be relatively good. The average knowledge score for Dengue and Chikungunya together was 5.4 ± 1.96 (SD), with a theoretical maximum of 9.

**Table 2:** Distribution of knowledge indicators by gender.

| **Table 2: Distribution of knowledge indicators by gender** |  |  |  |
| --- | --- | --- | --- |
| **Items accurately identified as correct** | **Total (n=309)**<br>**(Correct answers)** | **Men (n=154)**<br>**n (%)** | **Women (n=155)**<br>**n (%)** |
| **Symptoms related to Dengue** |  |  |  |
| High Fever | 235 (76) | 114 (74) | 121 (78) |
| Headache | 64 (21) | 36 (23) | 28 (18) |
| Pain behind eyes | 55 (18) | 28 (18) | 27 (17) |
| Joint pain | 88 (29) | 55 (36) | 33 (21) |
| Muscle and/or bone pain | 53 (17) | 31 (20) | 22 (14) |
| Rash | 57 (18) | 18 (12) | 39 (25) |
| Nausea and vomiting | 104 (34) | 49 (32) | 55 (35) |
| **Symptoms related to Chikungunya** |  |  |  |
| High Fever | 100 (32) | 49 (32) | 51 (33) |
| Headache | 31 (10) | 15 (10) | 16 (10) |
| Nausea and vomiting | 51 (17) | 27 (17) | 24 (15) |
| Joint swelling | 200 (65) | 96 (62) | 104 (67) |
| Muscle pain | 97 (31) | 53 (34) | 44 (28) |
| **Cause of Dengue** |  |  |  |
| Virus | 136 (44) | 72 (47) | 64 (41) |
| **Cause of Chikungunya** |  |  |  |
| Virus | 193 (66) | 101 (66) | 92 (59) |
| **Mode of transmission of Dengue** |  |  |  |
| Through mosquito bite | 215 (70) | 101 (66) | 114 (74) |
| **Mode of transmission of Chikungunya** |  |  |  |
| Through mosquito bite | 240 (78) | 117 (76) | 123(79) |
| **Prevention methods for Dengue** |  |  |  |
| Bed nets | 96 (31) | 45 (29) | 51 (33) |
| Indoor residual spray | 219 (71) | 112 (73) | 107 (69) |
| Prevent standing water near house | 250 (81) | 128 (83) | 122 (79) |
| Applying mosquito repellent on body | 98 (32) | 46 (30) | 52 (34) |
| **General Prevention methods for Chikungunya** |  |  |  |
| Similar to Dengue | 278 (90) | 143 (93) | 135 (87) |

Table 3 shows the respondents awareness about Dengue and Chikungunya. More than three-fourth of the respondents (90%) reported that they had previously heard about Dengue. When asked about availability of supportive and symptomatic treatment, 89% and 80% reported that such supportive and symptomatic treatment is available for Dengue and Chikungunya respectively. There was no statistically significant gender-differences across awareness indicators of Dengue and Chikungunya. Respondents awareness about Dengue and Chikungunya was found to be relatively good. The average awareness score for Dengue and Chikungunya together was 1.68 ± 0.63 (SD), with a maximum of 2.

**Table 3:** Distribution of awareness indicators by gender.

| **Table 3: Distribution of awareness indicators by gender** |  |  |  |
| --- | --- | --- | --- |
| **Items accurately identified as correct** | **Total (n=309)**<br>**(Correct answers)** | **Men**<br>**(n=154)**<br>**n (%)** | **Women**<br>**(n=155)**<br>**n (%)** |
| **Heard about Dengue or not** |  |  |  |
| Yes | 293 (90) | 151 (98) | 142 (92) |
| **Availability of supportive and symptomatic treatment for Dengue** |  |  |  |
| Yes | 275 (89) | 139 (90) | 136 (88) |
| **Availability of supportive and symptomatic treatment for Chikungunya** |  |  |  |
| Yes | 246 (80) | 123 (80) | 123 (79) |

The following figures 1 and 2 shows gender differences across the respondents correct knowledge and awareness for Dengue and Chikungunya. Although not statistically significant, in case of Dengue women have had better knowledge while men have had slightly better awareness with regard to Dengue. In case of Chikungunya, men respondents have had better knowledge and awareness as compared to women respondents, however, difference was not statistically significant.

**Figure 1.**
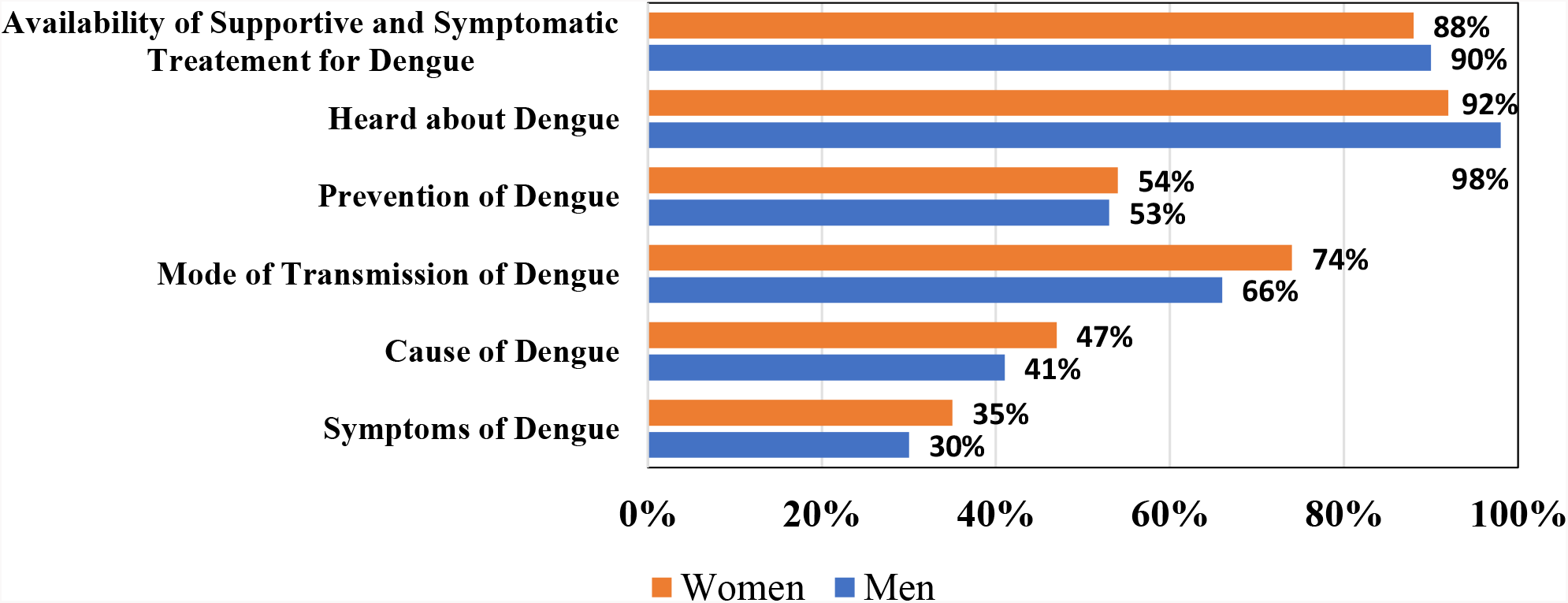
Proportion of maximal mean knowledge score (%) by topic as percentage of maximal score for Dengue by gender

**Figure 2.**
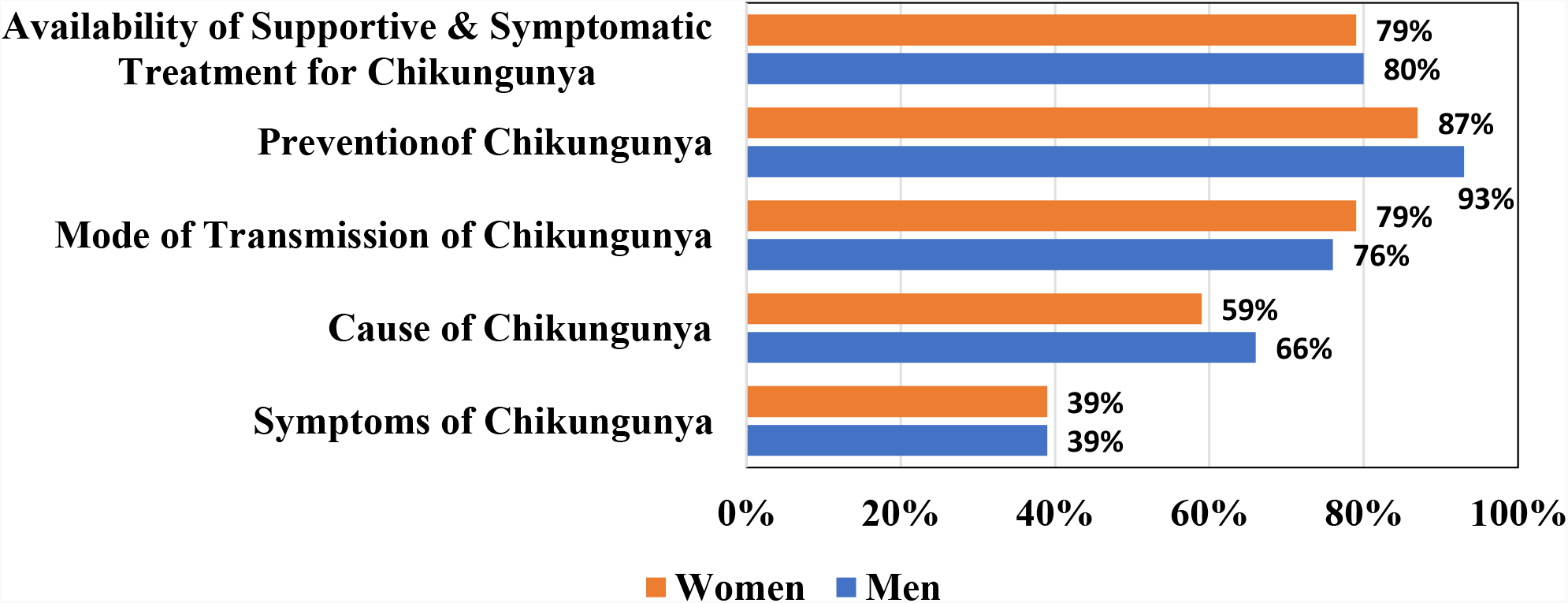
Proportion of maximal mean knowledge score (%) by topic as percentage of maximal score for Chikungunya by gender

Table 4 shows literacy status-wise differences about Dengue and Chikungunya across knowledge and awareness indicators. The t-test results suggest that there is a significant statistical difference in knowledge and awareness about symptoms, cause, prevention methods, mode of transmission and availability of treatment. Table 4 clearly shows that literate respondents had more knowledge and awareness about Dengue and Chikungunya than their illiterate counterparts.

**Table 4:** Literacy status-wise differences across knowledge and awareness indicators of Dengue and Chikungunya.

|  |  |  |  |  |  |  |
| --- | --- | --- | --- | --- | --- | --- |
| **Parameters** | **Illiterate n (%)** | **Literate n (%)** | **t-test** | **df** | **Significance (2-tailed)** | **P value** |
| **Knowledge** |  |  |  |  |  |  |
| Symptoms of Dengue and Chikungunya | 22 (76) | 246 (88) | 22.79 | 308 | 0.00 | < 0.001* |
| Causes of Dengue and Chikungunya | 9 (31) | 207 (74) | 26.74 | 308 | 0.00 | < 0.001* |
| Prevention & transmission of Dengue and Chikungunya | 28 (96) | 272 (97) | 101.32 | 308 | 0.00 | < 0.001* |
| **Awareness** |  |  |  |  |  |  |
| Availability of supportive and symptomatic treatment for Dengue and Chikungunya | 18 (62) | 263 (94) | 55.59 | 308 | 0.00 | < 0.001* |

\* Highly Significant df = degree of freedom

## Discussion

Against the background of the occurrence of recurrent outbreaks of Dengue and Chikungunya in Pune city in recent years^12^, present cross-sectional study documented the knowledge and awareness of general community members residing in low-income settings regarded as slums in Pune city with regard to Dengue and Chikungunya. Public awareness is considered a key to successful vector control programmes^4^. Such awareness is crucial as it improves people’s understanding of risks and of prevention strategies and is considered a key to success in vector-borne diseases control programmes^2,4^.

The study findings documented relatively better Dengue-related knowledge and awareness among the general population residing in slum areas of study amidst recurrent outrbeaks. Similar findings related to awareness about Dengue was also documented in a study conducted in urban low-income areas of south Delhi^8^. Knowledge about Dengue was comparatively more in women respondents while awareness was found to be comparatively more among the men respondents. The literate respondents had better knowledge and awareness about Dengue as compared to illiterate respondents. Similar findings have been reported by a study conducted in Karachi, Pakistan^13^. High fever was regarded as the most commonly reported symptom of Dengue. Similar findings have been reported by other studies conducted in different cities of India^9,14^. Knowledge about causes of Dengue was relatively low, as only 44% of respondents could report virus as a cause of Dengue. Most of the respondents reported mosquito bite as the mode of transmission of Dengue. Similar findings have been reported by a study conducted in Myanmar^15^. Stagnant water management was most commonly reported preventive method for Dengue. Most of the respondents in this study had previously heard information about Dengue. Similar findings have been reported by a study conducted in urban areas of south Delhi^8^. Most of the respondents in the study were aware about the availability of supportive and or symptomatic treatment for Dengue. The study findings also indicated that there was relatively better Chikungunya-related knowledge and awareness. Knowledge about Chikungunya was more among men respondents while awareness was almost identical in both men and women respondents. The literate respondents had better knowledge and awareness about Chikungunya as compared to illiterate respondents. Joint swelling was the most commonly reported symptom of Chikungunya. Knowledge about cause of Chikungunya was relatively high, and majority respondents could report virus as a cause of Chikungunya. Most of the respondents reported mosquito bite as the mode of transmission of Chikungunya. Similar findings have been reported by a study conducted in Dhaka city of Bangladesh^16^. Most of the respondents reported that prevention methods for both Dengue and Chikungunya are similar.

Most of the respondents were unable to differentiate and distinguish symptoms of VBDs. They mentioned few general symptoms of febrile illnesses. Also, respondents had comparatively less information and knowledge about causative agents of the Dengue and Chikungunya. These finding would feed and enable strengthening of the to be prepared National plan for effective community engagement and mobilization in vector control.

The World Health Organization GVCR strategy envisage that for achieving sustainable impact in vector control, increased intersectoral and interdisciplinary action is required, thereby linking and converging efforts in environmental management, health education, and reorienting relevant governments programmes around proactive strategies to control new and emerging threats^1^. Further the GVCR strategy already acknowledged the importance of engaging local authorities and communities wherein broad-based and inclusive intersectoral collaboration has been considered as important element for improved vector control delivery, through catering of interventions to specific local needs and scenarios as informed by locally generated epidemiological data^1^. In that regard the present study findings would help in planning and implementation of community mobilization strategies amidst recurrent outbreaks to facilitate community engagement and collaboration leading to development of sustainable control programme efforts.

## Conclusion

The findings from the study revealed that there were better knowledge and awareness levels about Dengue and Chikungunya in the urban slum population. These could be due to public health campaigns held during outbreak situations. The knowledge and awareness levels of VBDs were found to be more in literate respondents as compared to illiterates. This indicates urgent need of sustaining these knowledge and awareness into efficacious prevention practices to facilitate appropriate and sustained public health actions and individual risk protection measures.

## Programmatic recommendations

As revealed through the study, most of the respondents were unable to differentiate and distinguish symptoms of VBDs resulting into reporting of general symptoms of febrile illnesses. In view of the overlap of symptoms which have implications on early help-seeking for diagnosis and treatment specifically during the outbreak situations; differentiating features of Dengue and Chikungunya should be mentioned in the currently used NVBDCP health education materials so that the general community will be able to accurately differentiate, distinguish and could take necessary actions to prevent and control the VBDs and thereby recurrent epidemics. Further, it was also found that respondents had comparatively less information and knowledge about causative agent of the Dengue and Chikungunya. This suggests the need to incorporate more scientific aspects and information about the causative agents of VBDs in the health education materials used by the NVBDCP local programme authorities. There is overall need for aggressive health campaigning and rigorous health education activities in the low-income settings of urban communities particularly amidst outbreak situations, through effective health education strategies to promote successful and sustained implementation of WHO recommended GVCR strategy.

## Data Availability

SPSS data entry files will be uploaded, if requested by the editorial team.

## Key Messages

Findings indicate capitalizing on better knowledge and awareness levels achieved due to public health campaigns amidst recurrent epidemic/outbreak situations to facilitate appropriate public health actions on continual basis for strengthening vector control response and suggests need for aggressive and sustained health education campaigning to promote implementation of WHO recommended GVCR strategy.

## Authorship declaration

Swapnil Godbharle and Bibek Raj Giri conceptualized the study, prepared the study tools and participated in the data collection, analysed the data and wrote the manuscript. Abhay Kudale participated in conceptualization of the study, guided the design and coordination of the study, help analysed the data, and critically revised and reviewed the paper. All authors read and approved the final paper.

## Ethical approval

The study was approved by institutional ethics committee of Savitribai Phule Pune University.

## Acknowledgement

We gratefully acknowledge our institute the Interdisciplinary School of Health Sciences at Savitribai Phule Pune University for giving us an opportunity to conduct this study. Furthermore, we would like to thank Dr. Pradeep Awate, State Surveillance Programme Officer In-charge, Integrated Disease Surveillance Project, Maharashtra State for their facilitation and guidance in the initial planning stages of the study. Last but not least, we acknowledge the general community members from low-income settings of Pune city who participated in this study.

